# Proactive COVID-19 testing in a partially vaccinated population

**DOI:** 10.1101/2021.08.15.21262095

**Authors:** Ryan S. McGee, Julian R. Homburger, Hannah E. Williams, Carl T. Bergstrom, Alicia Y. Zhou

## Abstract

During the initial stages of the COVID-19 pandemic, many workplaces and universities implemented institution-wide proactive testing programs of all individuals, irrespective of symptoms. These measures have proven effective in mitigating outbreaks. As a greater fraction of the population becomes vaccinated, we need to understand what continued benefit, if any, proactive testing can contribute. Here, we address this problem with two distinct modeling approaches: a simple analytical model and a more simulation using the SEIRS+ platform. Both models indicate that proactive testing remains useful until a threshold level of vaccination is reached. This threshold depends on the transmissibility of the virus and the scope of other control measures in place. If a community is able to reach the threshold level of vaccination, testing can cease. Otherwise, continued testing will be an important component of disease control. Because it is usually difficult or impossible to precisely estimate key parameters such as the basic reproduction number for a specific workplace or other setting, our results are more useful for understanding general trends than for making precise quantitative predictions.

## 1 Introduction

The SARS-CoV-2 virus responsible for COVID-19 is particularly difficult to control because transmission often occurs in individuals who are pre-symptomatic or entirely asymptomatic (1–3). Throughout the COVID-19 pandemic, proactive testing has been a valuable tool for mitigating the spread of SARS-CoV-2 in workplaces, academic institutions, and other settings (4–7). The aim of proactive testing is to head off transmission from individuals who are not showing symptoms, by detecting infection early and isolating infected persons from the rest of the community.

Widespread COVID-19 vaccination is now underway in many parts of the world, ushering in the prospect of a full return to in-person work. As more individuals become vaccinated, can we afford to reduce or eliminate SARS-CoV-2 proactive testing from our disease control programs? Here we aim to address two aspects of this question. First, what level of vaccination and/or naturally-acquired immunity is required to render proactive testing programs unnecessary (8)? Second, as we transition from present conditions to that point, what are best practices for tapering off testing efforts?

## 2 Methods

To address the questions above, we use two distinct modeling approaches. First, we adapt a simple analytic approximation developed by Bergstrom et al. (9) to examine how testing and vaccination interact to reduce transmission. We illustrate these interactions as isoclines, or indifference curves, that indicate how increased vaccination coverage can compensate for reduced testing in a population. Second, we deploy the SEIRS+ modeling framework (10) used in our workplace testing (11) and return-to-school (7) models to consider workplaces or other groups in which vaccination efforts are underway. SEIRS+ is a stochastic, network-based epidemiological simulation model that accounts for the specific details of SARS-CoV-2 transmission and for the structure of social contact networks along which most infections are spread. We use the SEIRS+ model to simulate the dynamics of COVID-19 spread through a group of 1,000 individuals, following a single introduction from the community. We consider how the fraction of the population vaccinated and the extent of pre-existing natural immunity play into COVID-19 transmission dynamics.

### The exposure ratio model

To better understand the benefit of proactive SARS-CoV-2 testing in a partially vaccinated workplace or university setting, we adapt the Bergstrom et al. analytic model (9) to explore how vaccines and testing interact to reduce opportunities for COVID-19 transmission.

The model estimates how much the effective reproduction number *R*_*e*_—the average number of secondary cases generated by each primary case under current conditions—is reduced by a given level of vaccine uptake and a given cadence of proactive testing. While in practice this depends on the complex biology of the virus and the social dynamics in the host population, we can derive a simple approximation as follows. We envision that during each day of the infectious period, an infected individual is either “at-large” in the community or isolated at home. We assume that at-large individuals transmit at a constant rate corresponding to the effective reproduction number, while individuals isolated at home do not transmit. Testing serves to identify pre-symptomatic, asymptomatic, or paucisymptomatic individuals who would not have otherwise self-isolated. After testing positive, these individuals remove themselves from the population at large and isolate at home, which reduces the number of days they may expose others in the community. Suppose that testing the population every *n* days reduces the average amount of time that infectious individuals are at large by the *exposure ratio 𝒬*(*n*). The effective reproduction number is then reduced by a comparable fraction (Appendix A).

We now add vaccination and immunity from previous infection. Suppose that the basic reproductive number for the disease is *R*_0_, a fraction *γ* of the population is effectively vaccinated, and a fraction *η* is immune due to previous infection. If vaccination occurs independent of prior infectious status, the reproductive number with vaccination, testing, and previous immunity is given by

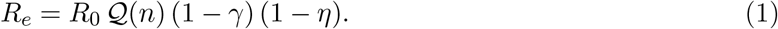

In words, the equation states that the effective reproduction number is equal to the product of the basic reproduction number, the fraction of exposure days that remain despite testing, the fraction of the population not yet effectively vaccinated, and the fraction of the population not yet naturally immune. In the Appendix A, we sketch out the analytic approach used to develop this equation, and provide a mathematical expression for the exposure ratio *𝒬*. Further details and a full derivation are provided in ref. (9).

### The SEIRS+ model

Our open source Python framework SEIRS+ implements stochastic network models of infectious disease transmission (https://github.com/ryansmcgee/seirsplus). This model extends the classic SEIRS model in a number of ways: it accounts for pre-symptomatic and asymptomatic disease states, it incorporates a process of quarantine or self-isolation, it accounts for individual heterogeneity in disease parameters, it allows an arbitrary distribution of residence times in each disease state, and it models transmission as occurring along a social contact network. We will briefly describe each of these aspects in turn. The SEIRS+ model has been used by a number of research groups (7, 12–14), and detailed documentation for can be found on the SEIRS+ github wiki (https://github.com/ryansmcgee/seirsplus/wiki).

Classic SEIR models of infectious disease are compartment models with compartments for susceptible (*S*), exposed (*E*), infectious (*I*), and removed (*R*) individuals. The SEIRS+ platform extends this framework to reflect the biology of SARS-CoV-2. In particular, the infectious class is divided into separate compartments for individuals who are in pre-symptomatic, asymptomatic, and symptomatic disease states. This allows us to model the decisions that some individuals make to self-isolate based on symptoms.

Because an important component of COVID-19 control involves isolating individuals in response to symptoms or testing, the SEIRS+ model introduces quarantine compartments that represent individuals in self-isolation (Figure 1). An individual may be quarantined in any disease state, and every disease state has a corresponding quarantine compartment. Quarantined individuals follow the same progression through the disease states, but their set of close contacts is defined by a reduced contact network. The transitions into quarantined states occur as the result of a positive test or self-diagnosis of symptoms. Individuals remain in the quarantine set of compartments until the designated isolation period is completed—ten days in this model. Thereafter they are moved into the non-quarantine compartment corresponding to their current disease state.

**Figure 1:**
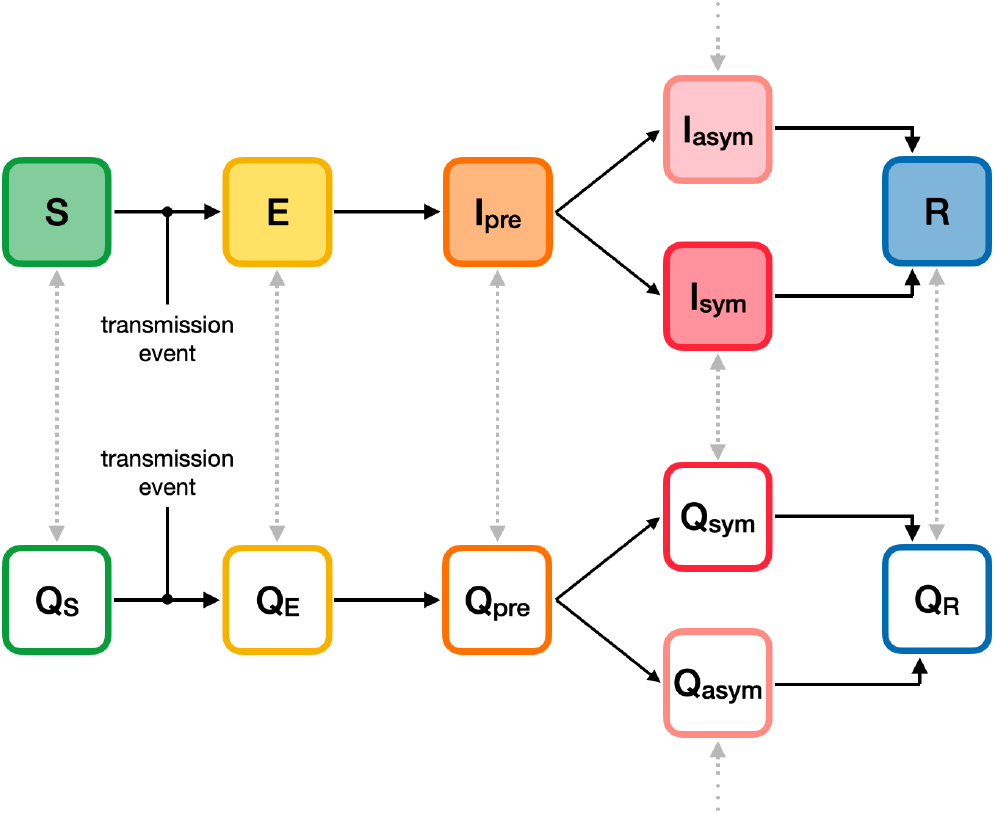
The SEIRS+ model extends the structure of a basic SEIR compartment model. In addition to the traditional *S, E*, and *R* states, the *I* state is split into three separate compartments: presymptomatic *I*_*pre*_, asymptomatic *I*_*asym*_, and symptomatic *I*_*sym*_. In addition, each primary compartment (shaded) has a parallel compartment *Q* (unshaded) representing individuals at the same disease stage but in quarantine.

In the SEIRS+ model, each individual in the model is independently assigned model parameters: for example, infectiousness, symptomaticity if infected, and the duration of time spent in each compartment if infected. This allows us to explicitly represent heterogeneity in disease characteristics (15, 16).

Traditional SEIR models assume that individuals transition among compartments at fixed rates, resulting in exponentially distributed residence times in each compartment. In reality, the distribution of residence times can be very different—and these differences can matter for modeling purposes (17, 18). The SEIRS+ model allows residence times drawn from gamma distributions based on empirical data. The dynamics governing disease transmission are proportional to contact rates between susceptible and infectious individuals, accounting for variation in infectiousness, and are described in detail in refs. (7) and (10).

### Contact networks

In their standard form, epidemic compartment models assume mass-action dynamics of disease transmission, equivalent to assuming that each individual in the population is equally likely to interact with any other individual. In practice, interactions are highly structured according the physical layout of workplaces as well as by the social contact networks that individuals inhabit (19, 20), and this can have important consequences for the dynamics of disease spread (21, 22). To account for this, the SEIRS+ model specifies a contact network of individuals who are prone to have “close contact” with one another. Those in one’s network might include family members, close friends, coworkers, roommates, romantic partners, and so forth. The model assumes that much—though not all—disease transmission occurs among the close contacts represented in the contact network. In the model, SARS-CoV-2 can also be transmitted among “casual” contacts, individuals who make infrequent, brief, and incidental contact such as might occur when passing through the same hallway or shopping in the same store. Here, we assume that 80% of transmission occurs along the network of close contacts, and the remaining 20% is casual in that it occurs at random among all members of the population (23).

The probability that a susceptible individual becomes infected depends on how susceptible they are to infection, how many of their contacts become infected, and how transmissible those infected contacts are. In a network transmission model, some individuals have numerous connections with others, and some have few. People with large numbers of contacts are unlikely to interact as closely with each one of them individually. To account for this, we assume a logarithmic rather than linear scaling of transmission opportunity as a function of network degree (7).

### Model parameters

In modeling the effects of testing and vaccination, we face multiple sources of uncertainty. Among these, the value of the basic reproduction number *R*_0_ looms largest. Under the traditional definition, *R*_0_ is the mean number of secondary infections generated by an index case in a susceptible population. For our purposes, the relevant value of *R*_0_ is the expected number of transmissions that occur within the institutional setting we are modeling—the number of transmissions that occur at work, for example. This value will depend on the physical structure of the workplace and the interaction patterns therein. Specifically, it will depend on the non-pharmaceutical interventions (NPIs) in place, including masking, ventilation, basic hygiene, and distancing procedures. And it will depend on the strains of SARS-CoV-2 circulating in the community. Because all of these factors vary from workplace to workplace and from week to week, obtaining a precise estimate of workplace *R*_0_ is seldom feasible. As such, the results of this model are more useful for understanding general trends than for making precise quantitative predictions.

The vaccines currently available in the US have demonstrated efficacy of around 65–95% against the wild-type form of SARS-CoV-2 (24–30). The most widely-distributed vaccines—the mRNA-based ones produced by Pfizer and Moderna—fall at the high end of that efficacy range. Thus far, these vaccines show comparable effectiveness against most circulating variants of concern as well (31–34). Available evidence increasingly suggests that these vaccines block SARS-CoV-2 symptoms and transmission at similar rates (35). Here we assume an average effectiveness of 90% against symptoms and transmission alike, intended to represent the mixture of vaccines available to the populations we are modeling. Appendix C shows model results when effectiveness is reduced to 70%.

## 3 Results

### Analytic approximation

Chance plays an important role in the dynamics of COVID-19 outbreaks. Even if two workplaces have very similar conditions, the introduction of an infected individual may seed a large outbreak in one and lead to no new cases in the other. But as a rule of thumb, an index case is unlikely to seed a sizeable and protracted outbreak when *R*_*e*_ *<* 1. Thus one prerequisite for safely operating a workplace is that mitigation measures therein are sufficient to drop *R*_*e*_ below unity.

To understand how non-pharmaceutical interventions, vaccination adoption, and proactive testing cadence contribute to reducing the likelihood of an outbreak, in Figure 2 we examine contour plots of the effective reproduction number *R*_*e*_ for three different *R*_0_ values. The horizontal axis in each panel represents the fraction of the population that have been vaccinated against COVID-19. The vertical axis indicates the cadence of proactive testing across the same population. Along each of the isoclines (solid black lines), the combined effect of proactive testing and vaccination is constant, i.e., each isocline corresponds to a fixed *R*_*e*_ value. Once *R*_*e*_ falls below unity, as indicated by the dashed line, substantial outbreaks are unlikely.

**Figure 2:**
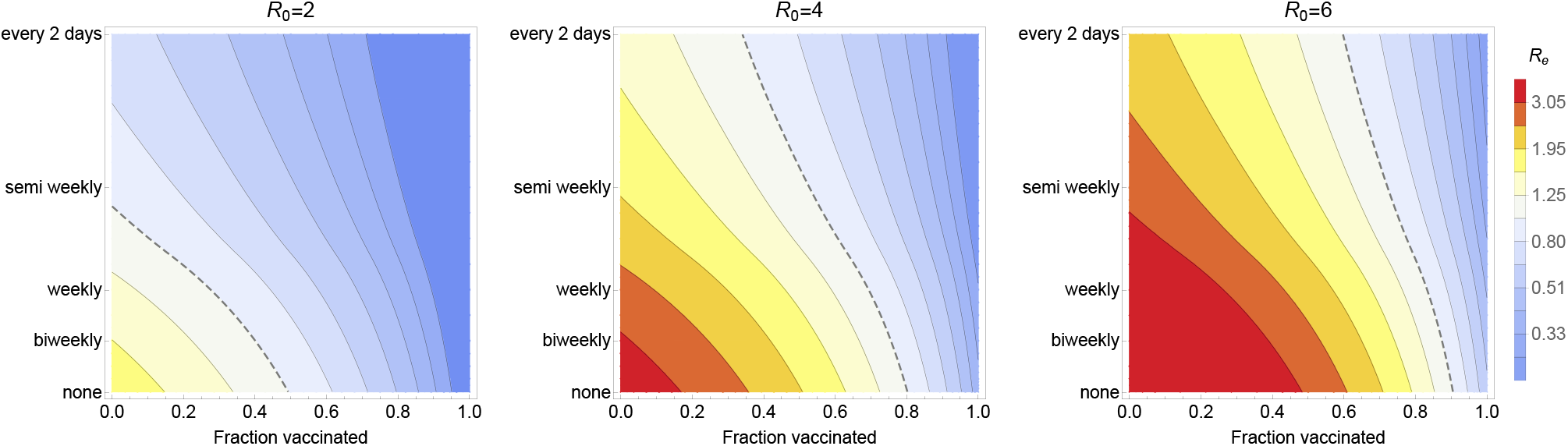
Effects of NPIs, vaccination, and testing. Contour plots of the effective reproduction number *R*_*e*_ show how mitigation efforts depend on NPIs via their effects on *R*_*e*_, on vaccination coverage (horizontal axis), and on testing cadence (vertical axis). The dashed line in each panel shows the combinations of vaccination and testing that are sufficient to drop *R*_*e*_ to unity for a given *R*_0_ value. (a) When *R*_0_ = 2.0, as we might expect for less transmissible variants with active NPIs, the effective reproduction number can be brought below unity with semi-weekly testing, by vaccinating half of the population, or some combination of those interventions. (b) With more transmissible variants and NPIs relaxed so that *R*_0_ rises to 4.0, broad vaccination coverage is an essential component to any control strategy and testing can help get down below the *R*_*e*_ = 1 threshold when vaccinate rates are in the range of 50-80%. (c) With highly transmissible variants and minimal NPIs, reducing *R*_*e*_ below one is impossible without very high levels of vaccination. In all three panels, we illustrate a situation in which 10% of the population has been previously infected and vaccines average 90% effectiveness. Additional parameters are summarized in Table B.2.

Panel 2a illustrates an *R*_0_ value of 2.0, reflecting a situation in which NPIs such as masks and distancing remain in place and in which the predominant SARS-CoV-2 strain is the original nonvariant virus. Panel 2b illustrates an *R*_0_ value of 4.0, as might be the case in a workplace where more transmissible variants are circulating yet NPIs have been relaxed. Panel 2c illustrates an *R*_0_ of 6.0, as we might see where the Delta strain is predominant and NPIs are limited or lacking entirely (36, 37). Taken together, these three plots illustrate the synergistic effects of NPIs, vaccination, and testing.

The more stringent the NPIs, the further the effective reproduction number *R*_*e*_ is reduced. Frequent testing and broad vaccination coverage also reduce *R*_*e*_. The higher the initial level of transmission, the greater the amount of testing and vaccination required to mitigate the risk of outbreaks.

When vaccine coverage is insufficient on its own to reduce the effective reproductive number *R*_*e*_ below one, a combination of proactive testing and NPIs can help considerably. For example, we see in Figure 2a that semiweekly testing has a comparable impact to vaccinating half the population. As vaccine coverage becomes more extensive, the effect of testing on *R*_*e*_ declines and eventually the benefit of testing becomes marginal.

In each of the three panels in Figure 2, *R*_0_ was fixed at a specific value and we looked at how a combination of proactive testing and vaccination reduces *R*_*e*_. Another way to look at the same mathematical expression is to fix the target level of *R*_*e*_ at 1, the level below which a sustained outbreak is unlikely, and then plot what value of *R*_0_ can be tolerated for a given vaccination level and testing rate (Figure 3). This graph allows a number of useful comparisons. For example, a vaccination rate of 60% might be enough to control the original SARS-CoV-2 strains even without supplementing with proactive testing. However, if more highly transmissible strains become established, driving the basic reproduction number upward toward 4 or even higher, vaccinating 60-70% of the population will not be sufficient to control disease on its own. Stronger NPIs and/or frequent, broad-scale proactive testing will be necessary.

**Figure 3:**
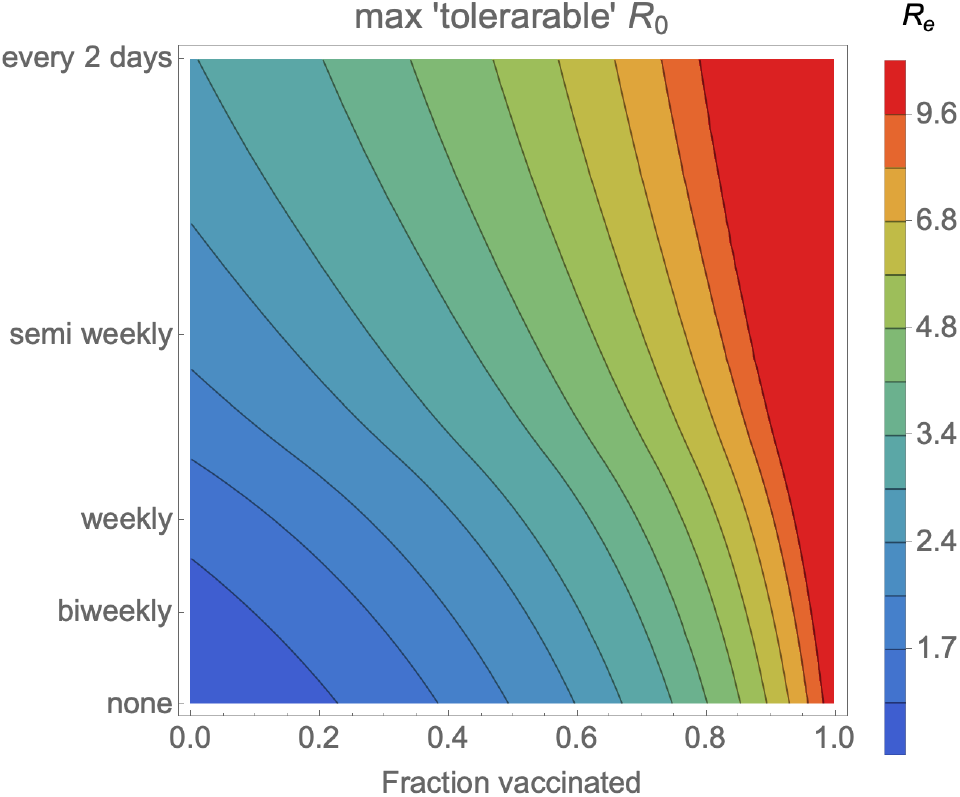
Maximum controllable *R*_0_ depends on vaccination and testing rates. Contour plot illustrates the highest *R*_0_ for which *R*_*e*_ can be brought down below 1.0 by means of the specified combination of vaccination coverage (horizontal axis) and testing cadence (vertical axis). Here 10% of the population has been previously infected and vaccines average 90% effectiveness.

### Stochastic network-based simulation

The analytical model above relies on a highly simplified picture of disease dynamics. To account for many of the complexities of the real world—superspreading, social contact networks, variation from person to person in disease progression, and the role of chance in disease outbreaks—we turn to the SEIRS+ simulation model.

Using this model, we consider the consequences of a single introduction into a workplace or other congregate setting of 1,000 individuals. Figure 4 illustrates the distribution of outbreak sizes resulting directly from this single introduction, at various testing cadences, as vaccine adoption increases. For each combination of parameters, we run 1,000 replicate simulations and depict the outbreak sizes as jitter plots where each dot represents the outcome of a single simulation run. The mean and 95th percentile outbreak sizes are indicated by the solid and dashed bars, respectively.

**Figure 4:**
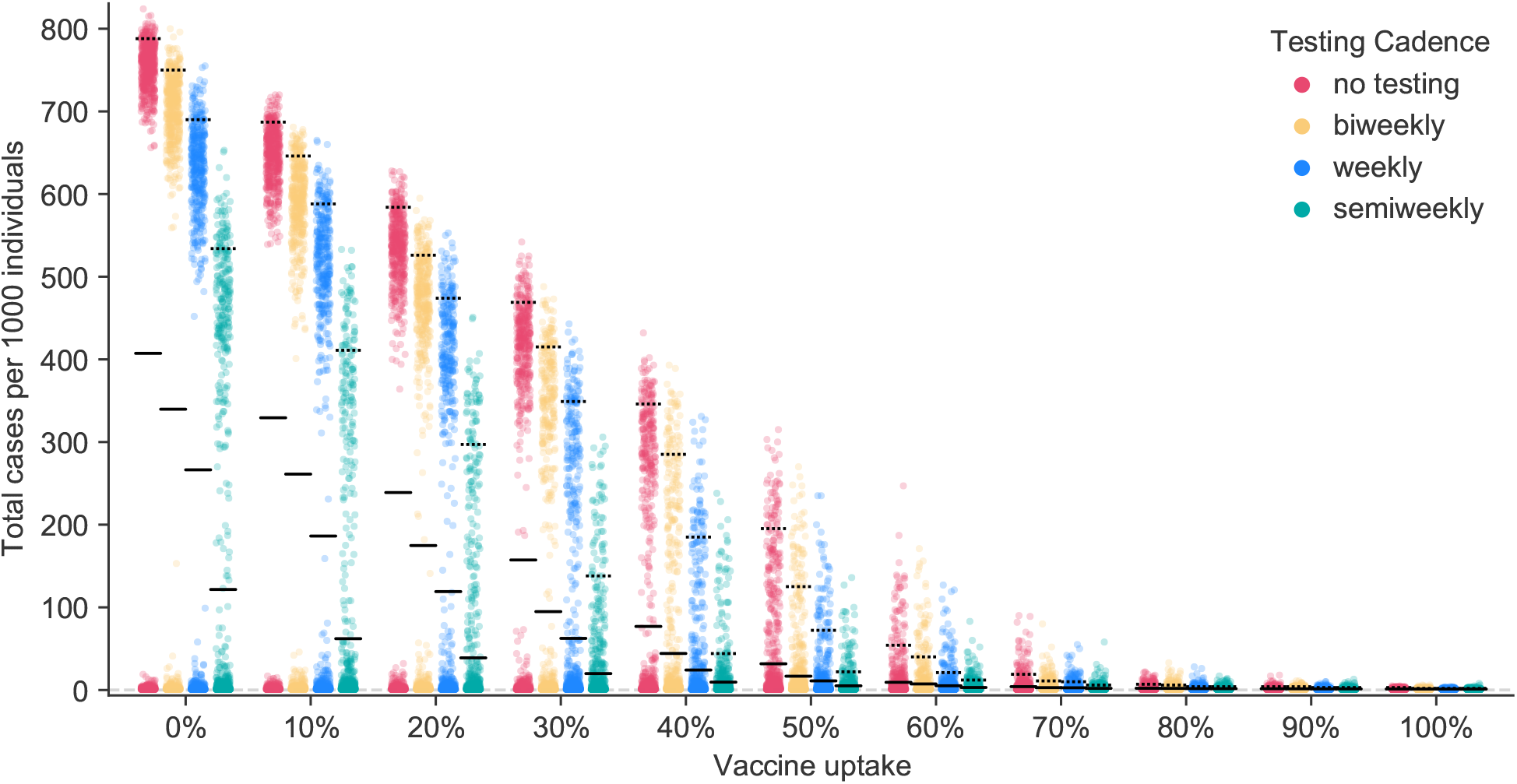
Outbreak sizes in the stochastic network simulation model. Here we illustrate the outcome of 1,000 simulations for each combination of testing cadence and vaccination uptake, when 10% of the population have previously been infected, vaccines are on average 90% effective at preventing infection and transmission, and *R*_0_ = 4.0. Each colored dot represents the outcome of a single simulation run. Solid black lines mark the mean outbreak sizes and dashed black lines mark the 95th percentile outbreak sizes for each parameter combination.

At lower levels of vaccine adoption and testing, we see a bimodal distribution of outcomes. In some simulation runs, large outbreaks occur, while in other runs with the same parameter settings, little or no transmission takes place at all. This is the consequence of chance events that contribute to the trajectory of disease spread. This figure also reveals that when less than two thirds of the population has been vaccinated, testing is a powerful tool for reducing both the mean number of cases and the 95th percentile outbreak size.

Figure 5 illustrates how the average benefit of testing declines as more of the population becomes vaccinated. The benefit of testing is measured as the reduction in the mean number of individuals infected after a single introduction. In the absence of vaccination and with *R*_0_ = 4.0, for example, weekly proactive testing prevents on average about 140 cases after a single introduction into a population of 1,000 people. By the time 70% of the population have been vaccinated, weekly testing prevents only a few cases. In general, we see that (1) when vaccination is limited, more frequent testing confers greater benefits, and (2) once vaccination becomes very common, the benefits of testing are substantially diminished.

**Figure 5:**
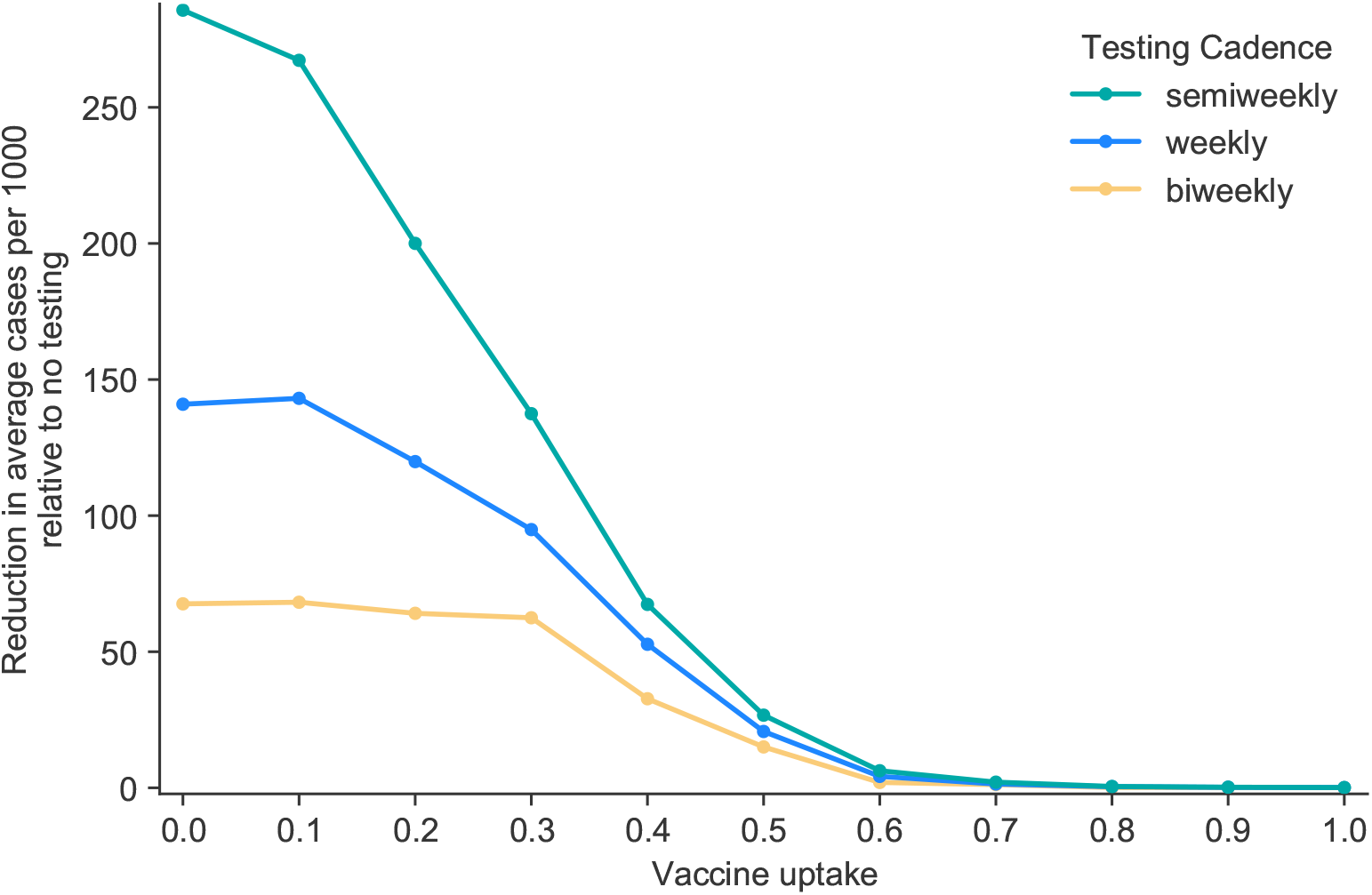
Value of testing in the stochastic network simulation model, as more of the population becomes vaccinated. Here we show how many cases per 1000 individuals are prevented on average when using a given testing cadence relative to not testing, where *R*_0_ = 4.0, 10% of the population have previously been infected, and vaccines are on average 90% effective at preventing infection and transmission.

Because Figures 4 and 5 illustrate the consequences of a single introduction, a combination of vaccine coverage and testing rate that poses an acceptable risk when community prevalence is low and introductions are rare may nonetheless be intolerably risky when community prevalence is high and frequent introductions are likely to occur.

Figures 4 and 5 illustrate the results of simulations for a single set of parameters: *R*_0_ = 4.0, prior incidence of disease 10%, and an average vaccine effectiveness of 90%. While these may be reasonable approximations of the situation in many locales, the general patterns that we observe are robust to changes in the precise parameter values. To illustrate, we have developed an interactive web application that displays results from the SEIRS+ model across a wide range of parameters (38). This application allows the user to select values of *R*_0_, vaccine efficacy, prior disease incidence, and testing cadence, and presents a jitter plot akin to Figure 4 for the chosen values.

## 4 Discussion

In this report, we have presented the results from two different modeling approaches: an analytic approximation and a stochastic network-based simulation. These approaches generate concordant results. Both indicate that testing is valuable at lower levels of vaccine adoption, regardless of *R*_0_. When *R*_0_ is high, either due to more transmissible strains or relaxation of non-pharmaceutical interventions, high levels of vaccination or a combination of vaccination and proactive testing will be necessary to achieve the *R*_*e*_ ≤ 1 threshold. Once the effective reproduction number *R*_*e*_ drops below 1 without testing, proactive testing offers little additional value and can be suspended if preventing outbreaks is the sole objective (see also ref. (8)).

This is not to say that proactive testing is useless in even more highly vaccinated populations. One-off transmission events are still possible even when the effective reproductive number *R*_*e*_ *<* 1 and a substantive outbreak is unlikely to occur. Proactive testing can reduce the probability of such transmissions occurring. Thus in addition to surveillance use, proactive testing can be helpful in situations where any transmission event whatsoever is considered unacceptable.

In previous modeling work, we found that testing generally offers a high return-on-investment in an unvaccinated workplace population (https://www.color.com/covid-19-outbreak-model). Here we find that the benefits of testing decline as vaccine adoption increases, but can be sizeable even in partially vaccinated cohorts.

In principle, one could adjust the testing cadence in real time, slowing the cadence as increasing fractions of the population become vaccinated. Given the inevitable uncertainties about the exact value of *R*_*e*_, and the logistical complexity of adjusting testing cadence in real time, it is likely to be difficult to fine-tune testing cadence by reducing the testing rate gradually as more people are vaccinated. As such, it may be logistically simpler to continue at testing at the original pre-vaccination testing cadence until there is good reason to believe that *R*_*e*_ *<* 1. At that point, the proactive testing program can be halted entirely.

## Supporting information

Mathematica code for analytical model

## Data Availability

The code for and data from the SEIRS+ simulations used in this paper are available at https://github.com/ryansmcgee/covid-testing-vaccinated-population
Mathematica code for the analytical model is included as supplementary material. The SEIRS+ codebase is open source and available at https://github.com/ryansmcgee/seirsplus.

## Acknowledgements

The authors thank Ted Bergstrom and Haoran Li for their part in developing the analytical model presented here. In part, this work was facilitated by the Hyak supercomputer system and funded by the STF at the University of Washington.

## Author contributions

Conceived of the models: CTB, RSM. Reviewed the literature: HEW, AYZ. Parameterized the models: CTB, RSM. Implemented the analytic model: CTB. Implemented the network-based model and ran the simulations: RSM. Analyzed the results: CTB, JRH, RSM, HEW, AYZ. Developed the interactive web app: JRH, HEW, AYZ. Drafted the manuscript: CTB, AYZ. Edited the manuscript: CTB, JRH, RSM, HEW, AYZ.

## Availability of data and material (data transparency)

The data from the SEIRS+ simulations are available at https://github.com/ryansmcgee/covid-testing-vaccinated-population

## Code availability

Mathematica code for the analytical model is included as supplementary material. The SEIRS+ codebase is open source and available at https://github.com/ryansmcgee/seirsplus. The specific code for the simulations here is available at https://github.com/ryansmcgee/covid-testing-vaccinated-population.

## A An analytic approximation for the effects of testing and vaccination

In the absence of vaccination, we define the exposure ratio *𝒬*(*n*) as the ratio of the mean exposure days when testing every *n* days, *Ē*(*n*), to the mean exposure days without any testing *E*ø:

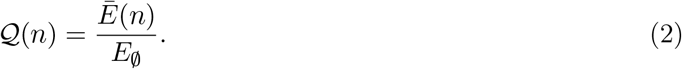

We calculate this quantity as follows. Even without testing, some individuals will choose to self-isolate after showing symptoms. If a fraction 1 − *u* of infected people develop symptoms and a fraction *v* of these isolate once symptomatic, the fraction who are symptomatic and choose to isolate is given by *v*(1 − *u*). The fraction of infected people who are asymptomatic or choose not to self-isolate is 1 − *v*(1 − *u*). Suppose that the presymptomatic infectious period is *y* days and the total infectious period is *C* days. In the absence of testing, the average number of exposure days is

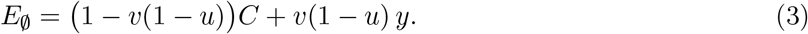

Let *E*(*t, n*) be the expected number of infectious days spent at large for a person who is infectious for *t* days when testing occurs every *n* days. For an individual who would not otherwise self-quarantine, testing reduces the number of infectious days at large from *C* to *E*(*C, n*). For an individual who would have self-quarantined anyway, testing can still pick up the infection before symptoms appear and thus reduces infectious days at large from *y* to *E*(*y, n*). The mean number of exposure days when testing every *t* days is then

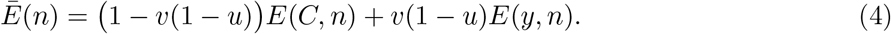

Now all we need is a way of calculating *E*(*t, n*). When the testing cadence is slower than the infectious period *C* minus the reporting delay *d* for the disease, an individual will be tested and results returned at most once during the course of infection. Bergstrom et al. (9) show that in this case, i.e. when *n > C* − *d*,

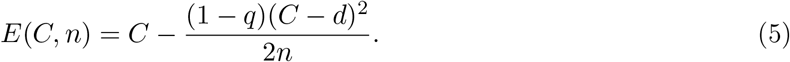

When an individual may be tested twice or more during the course of a single infection, the corresponding equation is somewhat more complicated. Denote the standard floor function by ⌊·⌋, let 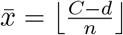, and let 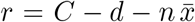. When the testing cadence is faster or equal to infectious period minus the reporting delay, *n* ≤ *C* − *d*,

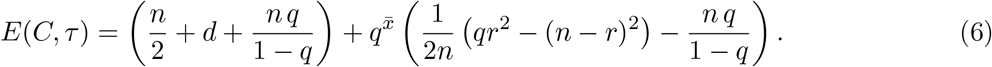

If infectiousness is constant throughout the duration of infection, testing every *n* days reduces the expected number of individuals infected by an index case by a fraction *𝒬*(*n*). Thus if the basic reproduction number without testing is *R*_0_, testing every *n* days would reduce this to 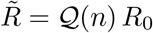

Next we incorporate vaccination and natural immunity. We assume that individuals who have been effectively vaccinated or who have recovered from previous COVID infection will not transmit COVID. If a fraction *γ* of the population has been effectively vaccinated, a fraction *η* have recovered from previous infection, and vaccination occurs independently of past infection status, the effective reproductive number with testing, vaccination, and previous infectious is given by

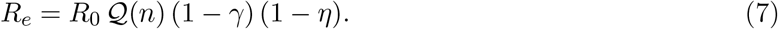

## B Overview of parameter values

Here we summarize the parameter values used in the analytic model (Table B.1) and in the SEIRS+ simulations (Tables B.2–B.4).

In the analytic model, we assume that test sensitivity is constant at 90% throughout the infectious period. In the SEIRS+ model we assume RNA-based testing with test sensitivities that change over the course of infection, as in ref. (7). Test sensitivity is 0% during the exposed period and non-infectious period, then climbs to 75% sensitivity for individuals in the first 2 days of their pre-symptomatic period and 80% sensitivity for any pre-symptomatic days beyond that. After the presymptomatic period ends, sensitivities for symptomatic and asymptomatic individuals alike follow the time course shown in Figure B1, with values based upon Levine-Tiefenbrun et al. (39). We assume there are no false positives.

**Figure B1:**
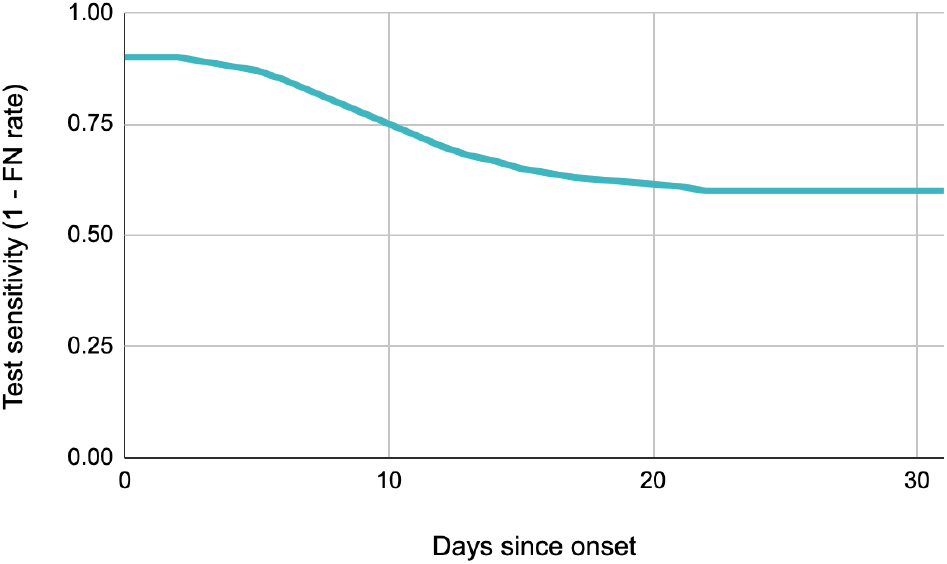
Test sensitivity in the SEIRS+ model. The probability of returning a positive test results when testing an infected, symptomatic individual as a function of the number of days since entering that state. The test sensitivity is equivalent to 1 minus the false negative rate.

**Table B.1.** Parameter values for the analytical approximation

| Parameter | Value or values |
| --- | --- |
| Basic reproduction number $R_0$ | 2.0, 4.0, and 6.0 as indicated in figure captions |
| Test sensitivity | 90% during the infectious period irrespective of symptomaticity. 0% prior to beginning of infectious period. |
| Delay between testing and quarantine | 1 day for those who test positive. |
| Infectious period | 6.2 days (irrespective of symptomaticity) |
| Presymptomatic period | 2.2 days (among those who eventually show symptoms) |
| Presymptomatic transmissibility | 100% relative to symptomatic transmissibility |
| Asymptomatic transmissibility | 100% relative to symptomatic transmissibility |
| Fraction of asymptomatic cases | 30% |
| Compliance with isolation upon symptoms | 30% |
| Compliance with testing | 100% |
| Vaccine effectiveness | 90% |
| Rate of disease with effective vaccine | 0% |
| Rate of transmission with effective vaccine | 0% |
| Fraction immune due to prior infection | 10% |

**Table B.2.** Overview of parameter values for the SEIRS+ model

| Parameter | Value or values |
| --- | --- |
| Basic reproduction number $R_0$<br>(mean individual reproduction number $R_0^{(i)}$) | 4.0 |
| Individual $R_0^{(i)}$ coefficient of variation | 2.0 (overdispersed) |
| Fraction of asymptomatic cases | 30% |
| Compliance with isolation upon symptoms | 30% |
| Compliance with testing | 100% |
| Disease state periods | See Table B.4 |
| Network structure | See Table B.3 |
| Proportion of global (off-network) transmission | 20% |
| Test sensitivity | Time-varying: see figure B1 |
| Vaccine effectiveness | 90% in main text, 70% in Appendix C. |
| Rate of disease with effective vaccine | 0% |
| Rate of transmission with effective vaccine | 0% |
| Fraction immune due to prior infection | 10% |

**Table B.3.**
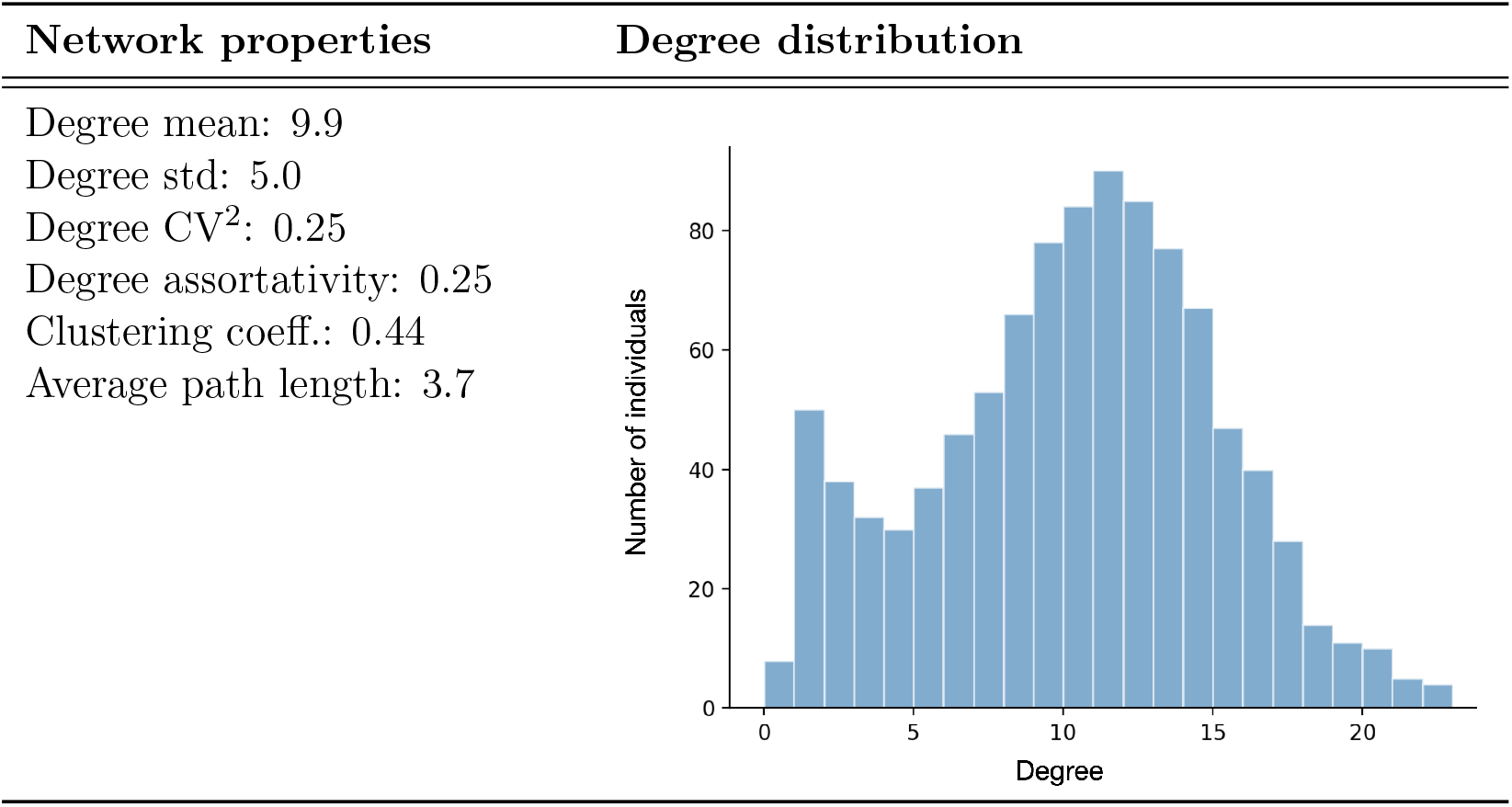
Network property statistics averaged across 1,000 replicate randomly generated contact networks and the degree distribution histogram for a representative random contact network.

**Table B.4.**
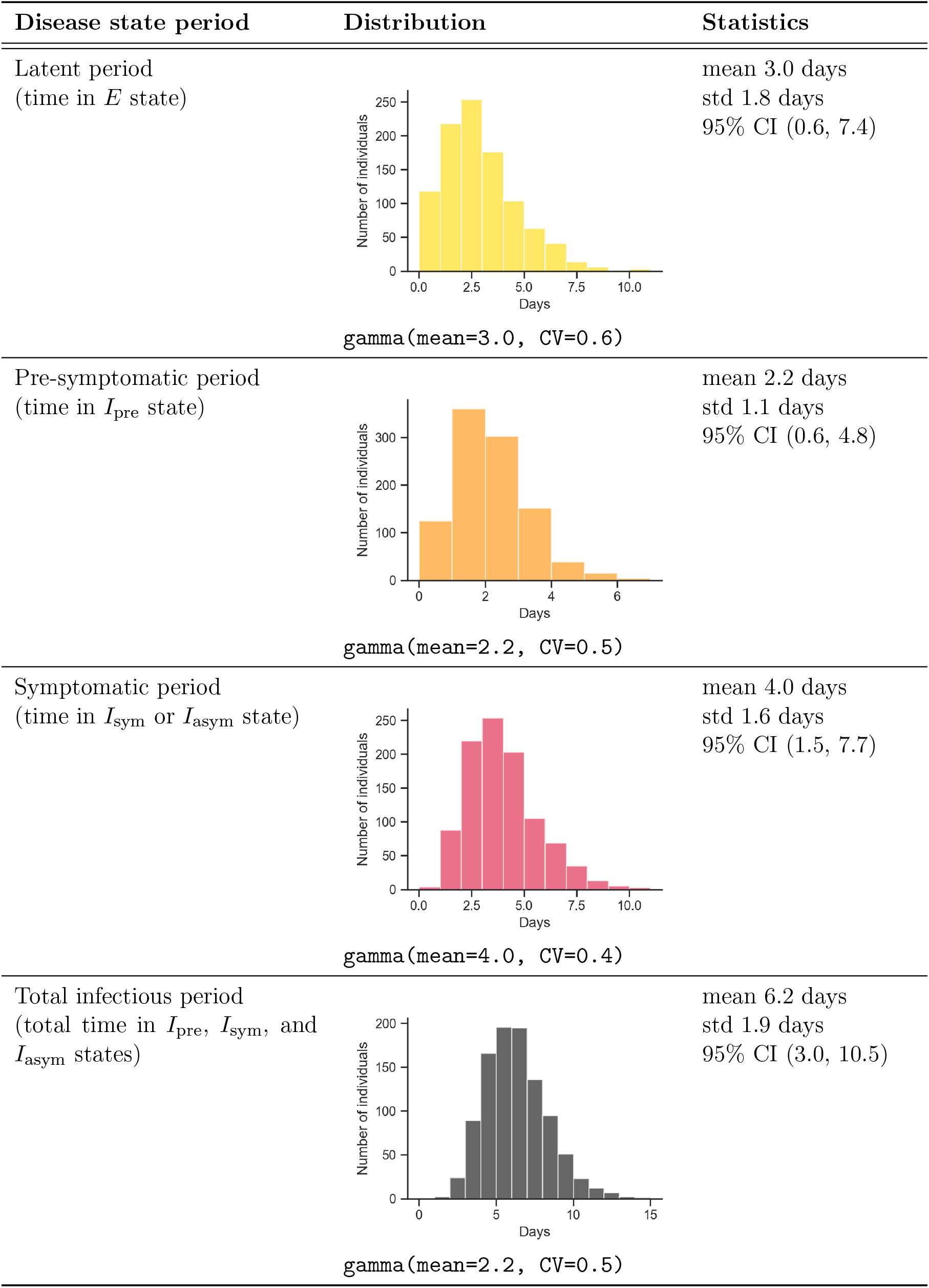
A representative distribution of period values drawn for population of 1,000 individuals is shown for each parameter in the center column below. Statistics across all replicate distributions in our analysis are shown in the rightmost column.

## C Lower vaccine efficacy

The SEIRS+ simulation results shown in Figures 4 and 5 of the main text are based on an average vaccine effectiveness of 90%. In some areas, the vaccines used may have lower effectiveness. To provide a sense of how that changes the results, here we show SEIRS+ simulation results for the case in which vaccine effectiveness is 70%. In this situation, testing remains valuable at higher vaccination fractions, but the overall patterns are qualitatively similar.

**Figure C1:**
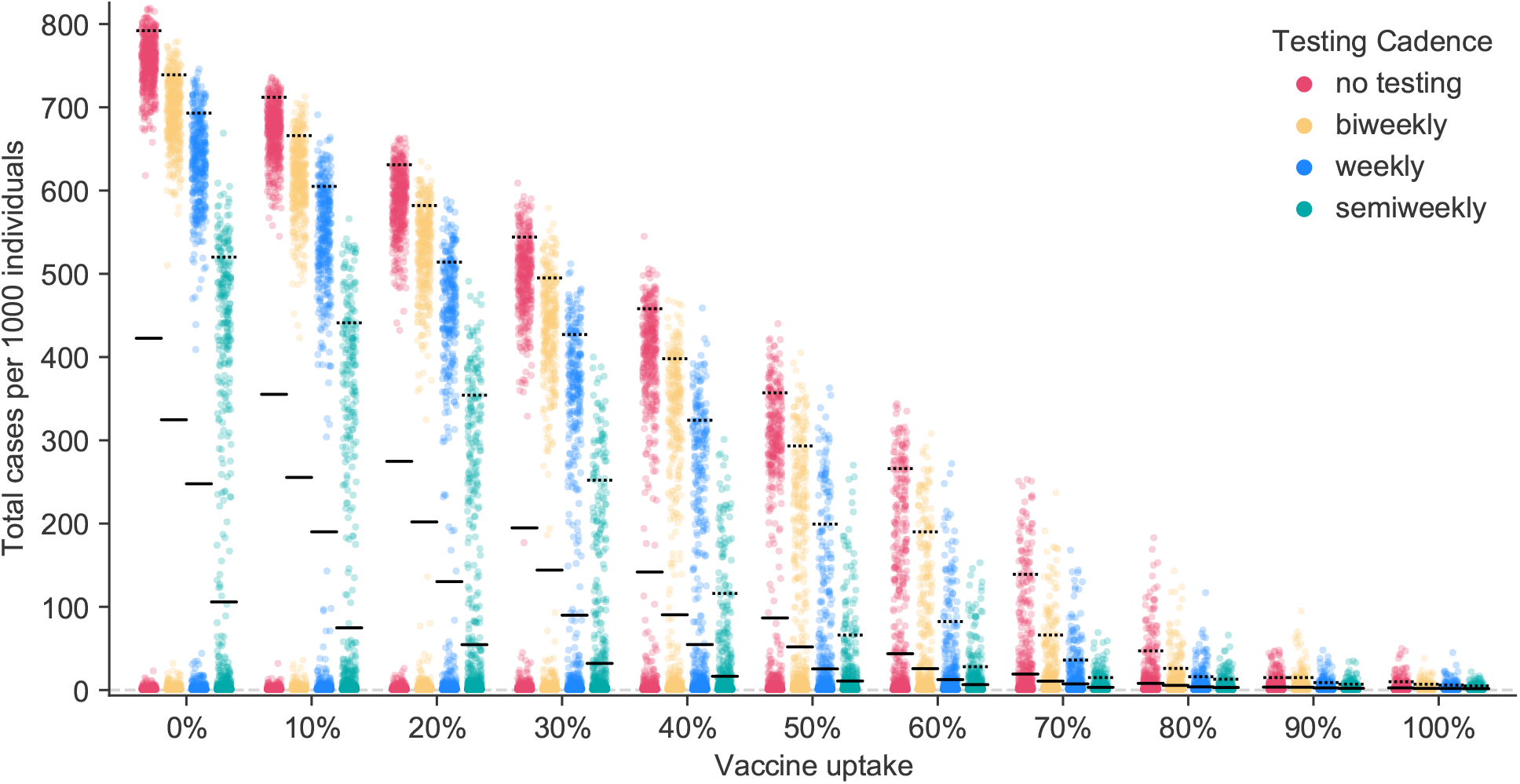
Outbreak sizes when vaccine effectiveness is 70%. Here we illustrate the outcome of 1000 simulations for each combination of testing cadence and vaccination uptake, when 10% of the population have previously been infected, vaccines are on average 70% effective at preventing infection and transmission, and *R*_0_ = 4.0. Solid black lines mark the mean outbreak sizes and dashed black lines mark the 95th percentile outbreak sizes for each parameter combination.

**Figure C2:**
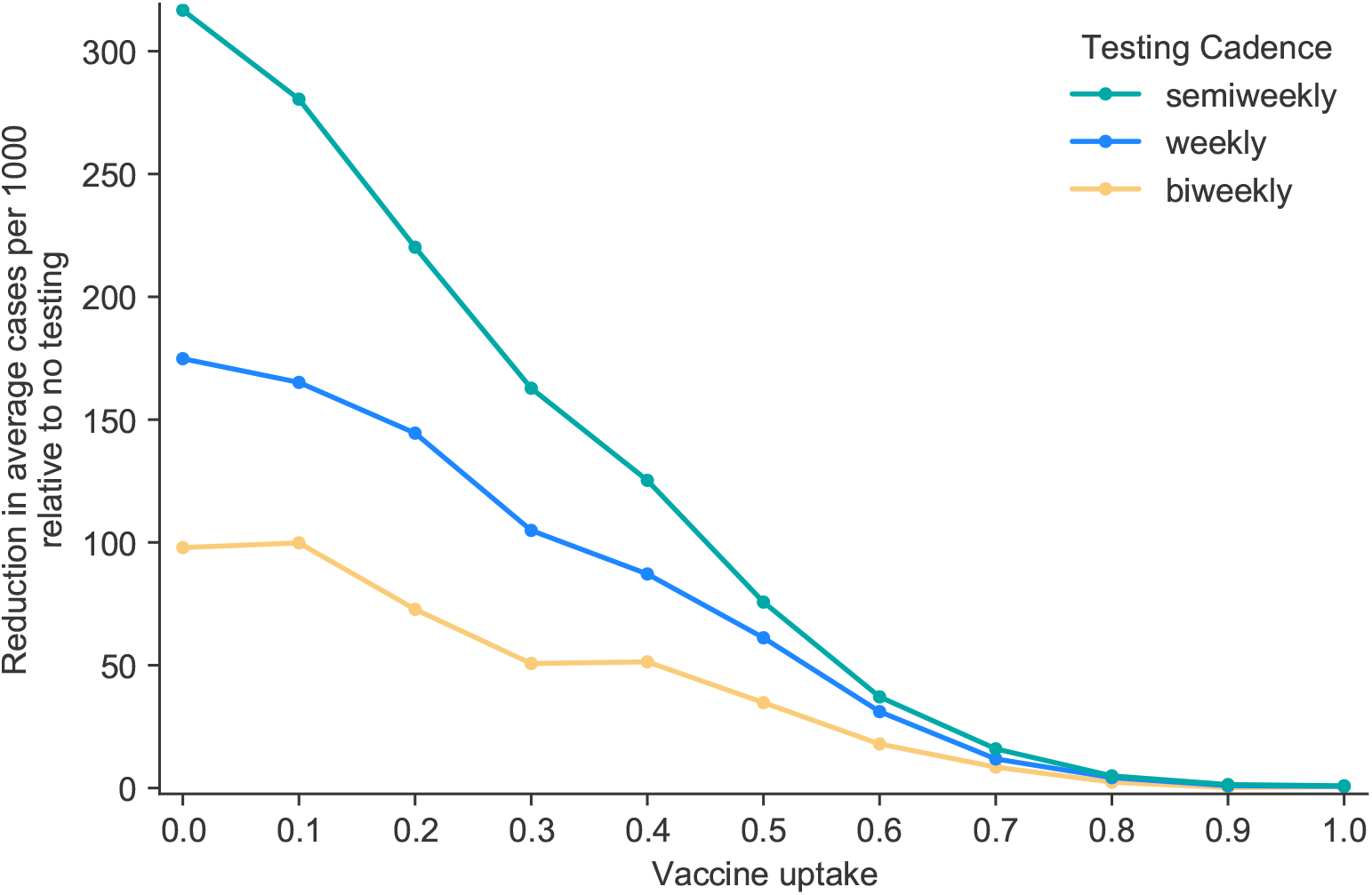
Value of testing when vaccine effectiveness is 70%. Here we show how many cases per 1000 individuals are prevented on average when using a given testing cadence relative to not testing, where *R*_0_ = 4.0, 10% of the population have previously been infected, and vaccines are on average 70% effective at preventing infection and transmission. As vaccination uptake increases, the benefits of testing here, with 70% effectiveness, decline less rapidly than the benefits with 90% effectiveness as shown in Figure 5. Differences between the values for 0% vaccine uptake here and those in Figure 5 are merely stochastic.

