## Supplementary material for "Proactive COVID-19 testing in a partially vaccinated population": Mathematica code for analytical model

#### *Mathematica* code for the analytical model

Ryan S. McGee, Julian R. Homburger, Hannah E. Williams, Carl T. Bergstrom, and Alicia Y. Zhou

#### Introduction

In McGee et al. 2021, we present two models, one analytical and one based on the SEIRS+ stochastic network simulation platform. This is the code for the analytical model.

We would like to know how testing, vaccination, and naturally acquired immunity interact in determining outbreak risk in a school or workplace. Our basic approach is to follow the method described in the *MedRxiv* preprint by Bergstrom, Bergstrom, and Li (2020), Frequency and accuracy of proactive testing for COVID-19. This model is used to compute the degree by which exposure days are reduced due to testing, in a simple deterministic model of disease course. We modify the model slightly to account for vaccination and immunity due to prior infection.

#### Model

##### Parameter values

We will assume that the infectious period is c=6.2 days.
Test sensitivity is 1-q=.9
The delay between testing and quarantine is d=1 day.
The fraction of asymptomatic cases is u=0.3
The fraction that self-isolate with symptoms is v=0.3
The presymptomatic period, in those who eventually develop symptoms, is 2.2 days.

In[1]:=
```
c = 6.2;
q = 0.1;
d = 1;
u = 0.3;
v = 0.3;
y = 2.2;
```

### Mathematical formulae

We implement the mathematical formulate from Bergstrom et al. 2020 *medRxiv* exactly. This allows us to calculate the exposure ratio to quantify the reduction in exposure days.

When testing occurs less than once per infectious period, the fractional reduction in infectious days is given by expression (2) in Bergstrom et al (2020).

In[7]:=
```
eLong[c_, {n_, q_, d_}] := c - ((1 - q) (c - d)^2)/(2 n)
```

$$\text{eLong}[c\_, \{n\_, q\_, d\_\}] := c - \frac{(1-q)\,(c-d)^2}{2\,n}$$

When testing occurs more than once per infectious period, the fractional reduction in infectious days is given by expression (5) in Bergstrom et al (2020).

In[8]:=
```
eShort[c_, {n_, q_, d_}] :=
 Module[
  {
   xbar = Floor[(c - d)/n],
   r = c - d - n Floor[(c - d)/n]},
  Piecewise[
   {{n/2 + d, q == 0}, {(n/2 + d + (n q)/(1 - q)) + q^xbar (1/(2 n) (q r^2 - (n - r)^2) - (n q)/(1 - q)), q > 0}}]]
```

$$\text{xbar} = \text{Floor}\left[\frac{c-d}{n}\right],\quad r = c - d - n\,\text{Floor}\left[\frac{c-d}{n}\right]$$

$$\left\{\left\{\frac{n}{2}+d,\ q==0\right\},\ \left\{\left(\frac{n}{2}+d+\frac{n\,q}{1-q}\right)+q^{\text{xbar}}\left(\frac{1}{2\,n}\left(q\,r^2-(n-r)^2\right)-\frac{n\,q}{1-q}\right),\ q>0\right\}\right\}$$

The function below is simply a convenient piece of code that selects the appropriate expression to use:

In[9]:=
```
eT[c_, {n_, q_, d_}] :=
 Piecewise[{{eLong[c, {n, q, d}], n > c - d}, {eShort[c, {n, q, d}], n <= c - d}}]
```

The mean number of exposure days under a testing regime is then given by equation (6) in Bergstrom et al (2020).

In[10]:=
```
e[{c_, u_, v_, y_}, {n_, q_, d_}] :=
 (1 - (1 - u) v ) eT[c, {n, q, d}] + (1 - u) v eT[y, {n, q, d}]
```

Without any testing at all, this simplifies to expression (7) in Bergstrom et al (2020).

In[11]:=
```
eNT[{c_, u_, v_, y_}] := (1 - (1 - u) v ) c + (1 - u) v y
```

When people in isolation do not transmit at all, we proceed by computing the exposure ratio (Defn. 4 in

Bergstrom et al 2020).

```
In[12]:= expRat[{c_, u_, v_, y_}, {n_, q_, d_}] := e[{c, u, v, y}, {n, q, d}] / eNT[{c, u, v, y}]
```

When there is no testing, we define the exposure ratio is 1. (Otherwise the equation above throws div zero errors).

```
In[13]:= expRat[{c, u, v, y}, {0, q, d}] := 1
```

Now we expand equation (8) in Bergstrom et al 2020 to define the new effective $R_e$ as the product of $R_0$, the fraction not yet effectively vaccinated (this rolls in both efficacy and reach), the fraction not yet in the removed category, and the exposure ratio from testing.

```
In[14]:= r[incidence_, effectivelyVaccinated_, r0_, {c_, u_, v_, y_}, {n_, q_, d_}] :=
          r0 (1 - effectivelyVaccinated) (1 - incidence) expRat[{c, u, v, y}, {n, q, d}]
```

### Visualization

#### Bar graph

We can then produce a bar graph illustrating $R_e$ values for various combinations of testing and vaccination. Below, with $R_0$=3 and 10% of the population immune due to previous infection:

In[15]:=
```
Show[{BarChart[
    Table[Map[r[.3, ev, 3, {c, u, v, y}, {#, q, d}] &, {0, 14, 7, 3.5}], {ev, 0, 1, 0.1}],
    ChartStyle → {ColorData[112, 1], ColorData[112, 3] , ColorData[112, 2],
      ColorData[112, 4]}, ChartLabels → {{"0", "0.1", "0.2", "0.3", "0.4",
       "0.5", "0.6", "0.7", "0.8", "0.9", "1.0"}, None}, BarSpacing → {0, 1},
    ChartLegends → {"Never", "Biweekly", "Weekly", "Semiweekly"},
    AxesLabel → {"effectively vaccinated", "R_e"}],
   Graphics[Line[{{1, 1}, {55, 1}}]]}, AspectRatio → 1 / 2,
  PlotRange → All, ImageSize → 1000,
  PlotLabel → "R_e=2.25"]
```

Out[15]=

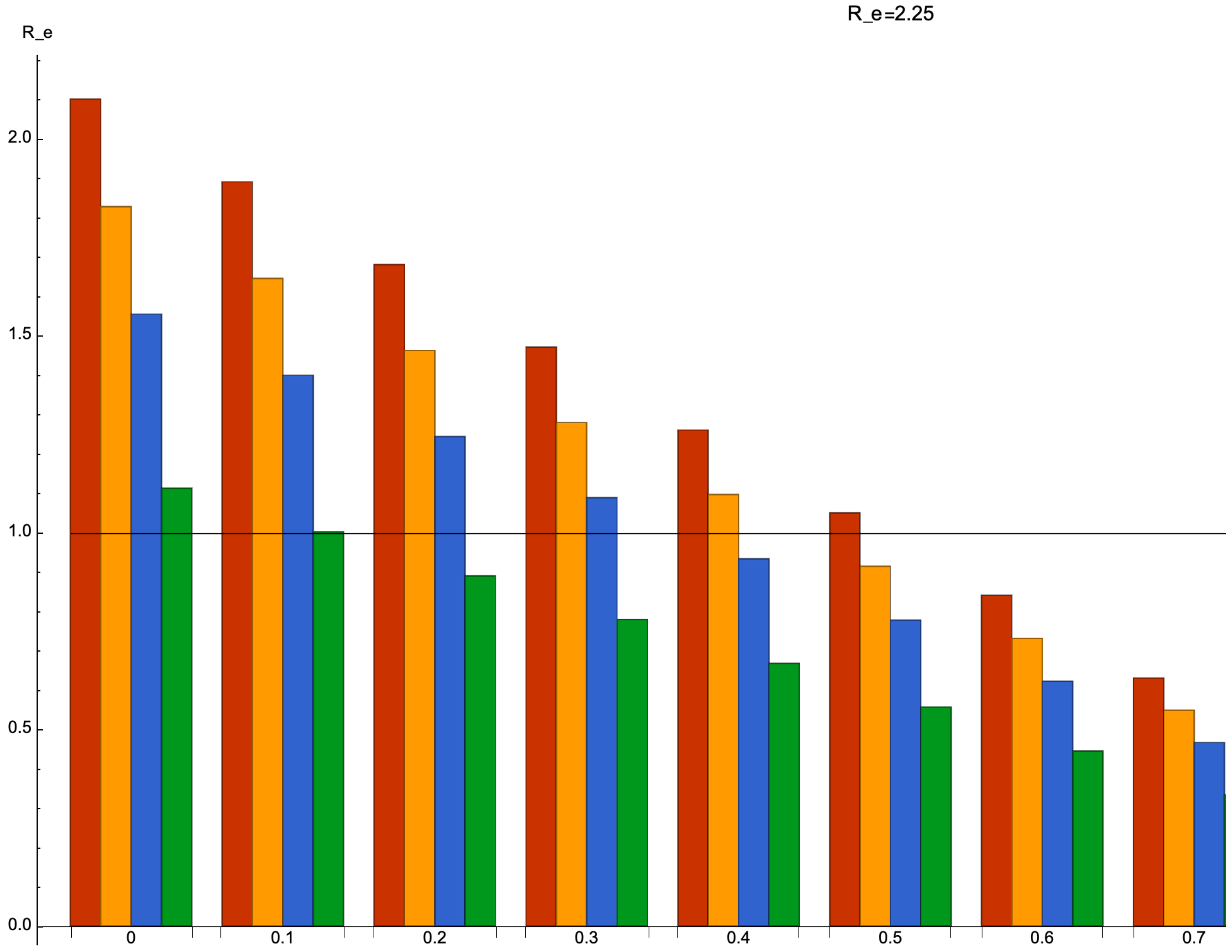


Note that because the terms of the expression for R influence one another multiplicatively, changing $R_0$ or the fraction already immune merely shifts the scale on the y axis (and thus raises or lowers the bar) without changing the relationship among heights across testing and vaccination rate classes.

#### Contour plot: $R_e$ due to vaccination and testing

Perhaps a more revealing way to plot these data is as a contour plot of indifference curves or isoclines. Because we have a closed-form expression for R, this is easy to do.

Suppose that we start with a population that is at 10%, and a 90% effective vaccine. Below, we plot $R_e$ values as a function of testing cadence and vaccination coverage for three different values of $R_0$.

In[16]:=
```
reContours = {
   {
    ContourPlot[
     r[.1, .9 * ev, 2, {c, u, v, y}, {1 / frequency, q, d}],
     {ev, 0, 1}, {frequency, .001, .5}, LabelStyle → 22,
     FrameLabel → {"Fraction vaccinated", ""}, RotateLabel → False,
     Exclusions → None,
     ColorFunction → ColorData[{"TemperatureMap", {-.5, 3}}],
     ColorFunctionScaling → False,
     Contours → Table[Round[.8^k, .01], {k, -5, 6}],
     ContourStyle →
      (If[.65 > # ≥ .50, {Directive[{Dashing[{.02, .01}], Thickness[0.005]}],
           {Directive[Black]}}] & /@ Table[Round[.81^k, .01], {k, -3, 6}]),
     FrameTicks → {{{{1 / 2, "every 2 days"}, {2 / 7, "semi weekly"}, {1 / 7, "weekly"},
         {1 / 14, "biweekly"}, {0, "none"}}, None}, {Automatic, None}},
     FrameLabel → {"Fraction vaccinated", ""}, ImageSize → 600,
     PlotLabel -> "R₀=2"],
    ContourPlot[
     r[.1, .9 * ev, 4, {c, u, v, y}, {1 / frequency, q, d}],
     {ev, 0, 1}, {frequency, .001, .5}, LabelStyle → 22,
     FrameLabel → {"Fraction vaccinated", ""}, RotateLabel → False,
     Exclusions → None,
     ColorFunction → ColorData[{"TemperatureMap", {-.5, 3}}],
     ColorFunctionScaling → False,
     Contours → Table[Round[.8^k, .01], {k, -5, 6}],
     ContourStyle →
      (If[.65 > # ≥ .50, {Directive[{Dashing[{.02, .01}], Thickness[0.005]}],
           {Directive[Black]}}] & /@ Table[Round[.81^k, .01], {k, -3, 6}]),
     FrameTicks → {{{{1 / 2, "every 2 days"}, {2 / 7, "semi weekly"}, {1 / 7, "weekly"},
         {1 / 14, "biweekly"}, {0, "none"}}, None}, {Automatic, None}},
     FrameLabel → {"Fraction vaccinated", ""}, ImageSize → 600,
     PlotLabel -> "R₀=4"],
    ContourPlot[
     r[.1, .9 * ev, 6, {c, u, v, y}, {1 / frequency, q, d}],
     {ev, 0, 1}, {frequency, .001, .5}, LabelStyle → 22,
     FrameLabel → {"Fraction vaccinated", ""}, RotateLabel → False,
     PlotLegends → Placed[
       BarLegend[Automatic, LegendMarkerSize → 400, LegendLabel → "Rₑ"], Right],
     Exclusions → None,
     ColorFunction → ColorData[{"TemperatureMap", {-.5, 3}}],
     ColorFunctionScaling → False,
```

```
    Contours → Table[Round[.8^k, .01], {k, -5, 6}],
    ContourStyle →
     (If[.65 > # ≥ .50, {Directive[{Dashing[{.02, .01}], Thickness[0.005]}],
          {Directive[Black]}}] & /@ Table[Round[.81^k, .01], {k, -3, 6}]),
    FrameTicks → {{{{1/2, "every 2 days"}, {2/7, "semi weekly"}, {1/7, "weekly"},
         {1/14, "biweekly"}, {0, "none"}}, None}, {Automatic, None}},
    FrameLabel → {"Fraction vaccinated", ""}, ImageSize → 600, PlotLabel -> "R₀=6"]
   }
  } // TableForm
```

Out[16]//TableForm=

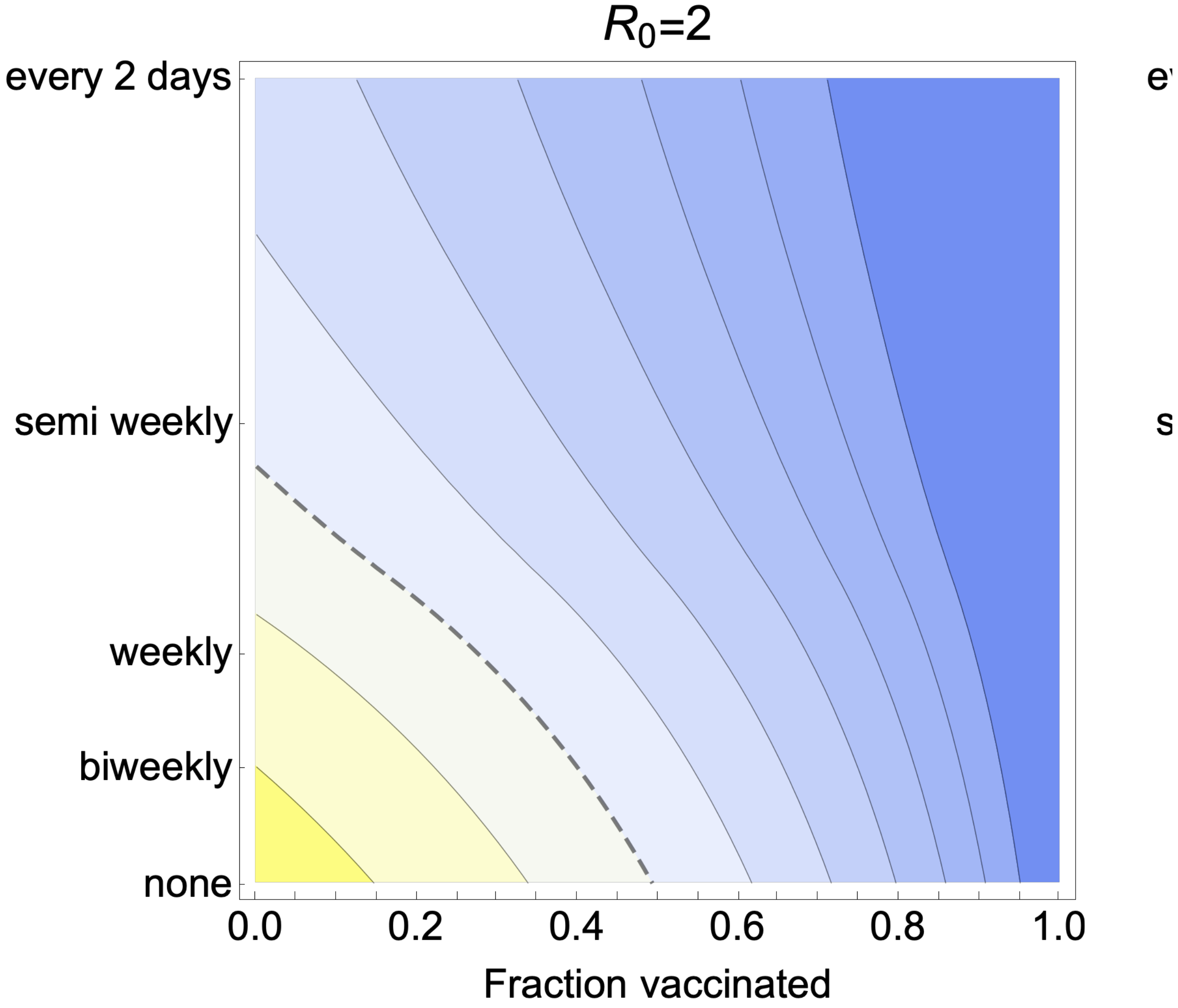


```
Export["Re.png", reContours];
Export["Re.pdf", reContours];
```

#### Contour plot: Maximum tolerable $R_0$

Alternatively, we could plot this in terms of the control needed to render the pandemic manageable

(Re<1) for a given initial $R_0$.

In[17]:=
```
tolerableR[incidence_, effectivelyVaccinated_, {c_, u_, v_, y_}, {n_, q_, d_}] :=
 1 / ((1 - effectivelyVaccinated) (1 - incidence) expRat[{c, u, v, y}, {n, q, d}])
```

In[18]:=
```
tolerableR0Plot =
 ContourPlot[tolerableR[.1, .9 * ev, {c, u, v, y}, {1 / frequency, q, d}], {ev, 0, 1},
  {frequency, .001, .5}, LabelStyle → 20, Exclusions → None, ImageSize → 600,
  ColorFunction → ColorData[{"Rainbow", {-1, 10}}], ColorFunctionScaling → False,
  Contours → Round[Table[1.19 ^ k, {k, 2, 13}], .1], PlotRange → All,
  FrameTicks → {{{{1 / 2, "every 2 days"}, {2 / 7, "semi weekly"}, {1 / 7, "weekly"},
      {1 / 14, "biweekly"}, {0, "none"}}, None}, {Automatic, None}},
  FrameLabel → {"Fraction vaccinated", ""}, PlotLegends →
   Placed[BarLegend[Automatic, LegendMarkerSize → 500, LegendLabel → "Re"], Right],
  ImageSize → 500, PlotLabel → "max 'tolerarable' R0"]
```

Out[18]=

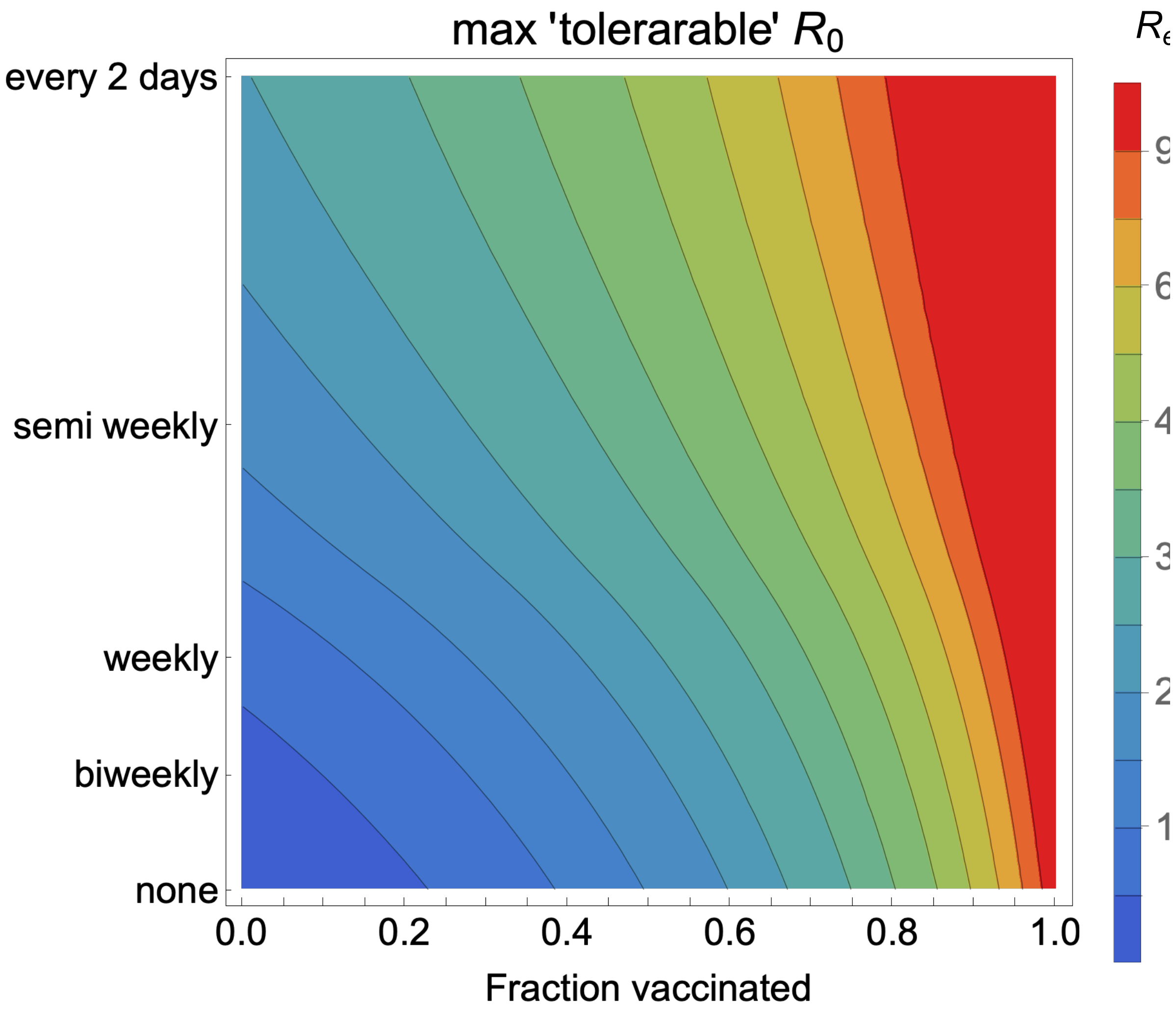


```
Export["tolerableR0.png", tolerableR0Plot];
```
